# Nuclear genetic modulation of tissue-specific mitochondrial RNA processing contributes to common disease risk

**DOI:** 10.1101/2025.08.22.25334213

**Authors:** Eleonora Centanini, Oliver Pain, Patrick F. Chinnery, Alan Hodgkinson

**Affiliations:** Department of Medical and Molecular Genetics, School of Basic and Medical Biosciences, King’s College London, London, SE1 9RT, UK; Maurice Wohl Clinical Neuroscience Institute, Department of Basic and Clinical Neuroscience, Institute of Psychiatry, Psychology and Neuroscience, King’s College London, London, United Kingdom; Department of Clinical Neuroscience & Medical Research Council Mitochondrial Biology Unit, School of Clinical Medicine, University of Cambridge, Cambridge CB2 0QQ, UK

## Abstract

Mitochondrial dysfunction is implicated in many common human diseases, yet it remains unclear whether it plays a primary causal role in pathogenesis or arises as a consequence disease-associated change. Moreover, the tissue-specific manifestation of mitochondrial disorders remains poorly understood, despite mitochondria being present in nearly all cell types. To address these questions, we leveraged over 15,000 RNA-sequencing datasets from 49 tissue types in the GTEx project, integrated with matched germline genetic data, to explore the causal impact of mitochondrial DNA (mtDNA) transcriptional processes on common disease risk. First, we identified 25 peak nuclear genetic variants associated with mtDNA transcript abundance, implicating nuclear genes including *FASTKD4*, *FASTKD5* and *PNPT1*, and revealing highly gene- and tissue-specific regulatory architectures. Next, we developed tissue-specific genetic scores to predict mtDNA-encoded transcript levels and validated their performance in independent datasets. Applying these scores to 377,439 individuals from the UK Biobank, we identified significant associations between predicted mtDNA transcript abundance and numerous common diseases and health-related quantitative traits, many showing marked tissue specificity. These included novel associations between hypertension and aorta-specific mitochondrial transcript abundance, and between Complex I gene expression and idiopathic Parkinson’s disease in neural crest-derived tissues, suggesting a causal role for mtDNA transcriptional processes in these conditions. Our results show that stable, genetically driven variation in mtDNA-encoded expression contributes to complex trait biology, and that nuclear-genetic regulation of mitochondrial RNA processing plays a key role in common disease susceptibility, rendering it a compelling therapeutic target.

## Main

Mitochondrial damage and dysfunction are observed in nearly all common diseases, particularly those linked to ageing. For example, increased mitochondrial DNA (mtDNA) damage^1^ and oxidative phosphorylation (OXPHOS) defects^2^ are reported in coronary artery disease, mitochondria-derived reactive oxygen species are implicated in diabetic complications^3^, metabolic reprogramming is central to cancer progression^4^ and dysfunctional mitochondria are a hallmark of many neurological disorders^5^. However, in many cases it remains unclear whether these mitochondrial changes play a primary causal role in disease pathogenesis or arise as secondary consequences of cellular stress.

Mitochondria possess their own compact genome, encoding just 13 protein-coding genes essential for the OXPHOS system that drives aerobic energy production, while ∼1100 nuclear-encoded proteins co-ordinate most mitochondrial functions, including the regulation of mtDNA replication, transcription, and translation. Fixed mutations in mtDNA (homoplasmies, affecting all mtDNA molecules) have been associated with diseases such as type 2 diabetes^6^, Parkinson’s disease^7^ and Alzheimer’s disease^8^. Additionally, large-scale studies have linked mtDNA variation to a broad range of disease-relevant traits, including insulin levels^9^, liver enzymes and kidney function^10^. While these findings suggest a potential causal role for mitochondrial processes in common disease, they are often difficult to replicate^11,12^ and yield conflicting results where the same mtDNA variant increases the risk of one disease but decreases the risk of another. These paradoxical findings are difficult to explain given that all are thought to influence OXPHOS, highlighting a limited understanding of the underlying mechanisms driving these relationships, and the complex regulatory interactions between the mitochondrial and nuclear genomes.

Importantly, most common disease-associated variants map to non-coding regions of the nuclear genome, implicating gene regulation as a key mechanism^13^. Yet, the genetic regulatory landscape of the mitochondrial genome, particularly the transcription and processing of mtDNA-encoded genes, remains poorly understood. Unlike nuclear genes, mtDNA is transcribed as multi-gene (polycistronic) precursors, processed by specialised machinery and regulated via extensive nuclear-mitochondrial crosstalk^14,15^. These processes vary across tissues and are increasingly implicated in disease^16,17^, but we currently lack a systems-level understanding of how genetically driven differences in mtDNA transcription and processing influence human health. Because mtDNA transcription is under both nuclear and mitochondrial genetic control^18–20^, it provides one window into lifelong, genetically programmed variation in mitochondrial activity, offering the opportunity to assess its role in disease, independent of downstream or reactive effects.

Here, we address this challenge by developing predictive genetic models of mtDNA-encoded transcript abundance across 49 tissues using RNA sequencing and whole-genome data from the Genotype-Tissue Expression (GTEx) project. We validate these models in independent datasets and apply them to >375,000 individuals in the UK Biobank to perform association testing across 699 diseases and 127 quantitative traits. In doing so, we uncover robust and biologically coherent links between mitochondrial transcriptional processes and common disease, providing new insight into the genetic and regulatory architecture of mitochondrial function in human health.

## Results

To characterise levels of mtDNA-encoded transcript abundance we considered RNA sequencing data from 15,066 samples across 49 tissue types from the GTEx project^21^. Data were reprocessed under stringent criteria to reduce the impact of misplaced reads (particularly those that may be influenced by nuclear encoded mitochondrial sequences, NUMTs) and transcript levels (Transcripts per million, TPM) were quantified for 13 protein-coding genes and two ribosomal RNAs encoded in the mitochondrial genome (mtDNA).

### Nuclear genetic control of mtDNA-encoded transcript abundance

To identify nuclear genetic variation associated with the transcript levels of genes encoded in mtDNA, we obtained whole genome sequencing data within the GTEx project for the same samples that we had RNA sequencing, and then performed association analyses between common genetic variation (minor allele frequency, MAF>5%) and the transcript levels of each of the 15 mtDNA-encoded genes separately within each tissue type, controlling for genetic ancestry and expression residuals (PEER factors^22^). For four tissue/cell types (whole blood, non-sun exposed skin, subcutaneous adipose and lymphoblastic cell lines (LCLs)) we then combined summary statistics from GTEx data with those obtained in independent datasets, as described in Ali *et al.* (2019)^18^.

Across all tissue types, we identify a total of 25 trans-genome expression QTLs (eQTLs, peak nuclear genetic variant-expression pairs) for mtDNA-encoded transcript abundance after Bonferroni correction (P<5x10^-8^/15 genes, table 1, sample sizes and inflation factors shown in supplementary table 1). To identify the potential nuclear gene through which each association is acting, we first considered whether any of the peak genetic variants are either functional (missense, nonsense or splice site variants), or in high linkage disequilibrium (R^2^>0.8) with a known functional variant. Doing so highlighted five genes that are linked to 21 out of the 25 associations (table 1), all of which have known roles in mitochondrial processes. *FASTKD4 (*example shown in Figure 1A) and *FASTKD5* are involved in the regulation of mitochondrial RNA processing and apoptosis; *MRPS35* encodes a mitochondrial ribosomal protein that is essential for protein synthesis within the mitochondria; *PNPT1* is responsible for the import of RNA into the mitochondria and plays a role in mitochondrial RNA processing; and *MTPAP* is involved in the polyadenylation of mitochondrial RNA, which is crucial for mitochondrial gene expression. Second, for the remaining four associations that were not linked with a functional variant, we performed mediation analysis, requiring a significant association between the peak genetic variant and a nearby nuclear gene (within 1MB, P<0.05), with the expression of that nuclear gene significantly mediating the effect of the genetic variant on the transcript abundance of the corresponding gene in mtDNA (P<0.05). Applying this approach, *KLHL41* (a gene that is not known to be involved in mitochondrial processes) was linked to the expression of *MTND3*, although we note that the top Open Targets^23^ gene (V2G score) for the peak nuclear genetic variant is *METTL5*, which is known to modify mtRNA.

**Figure 1:**
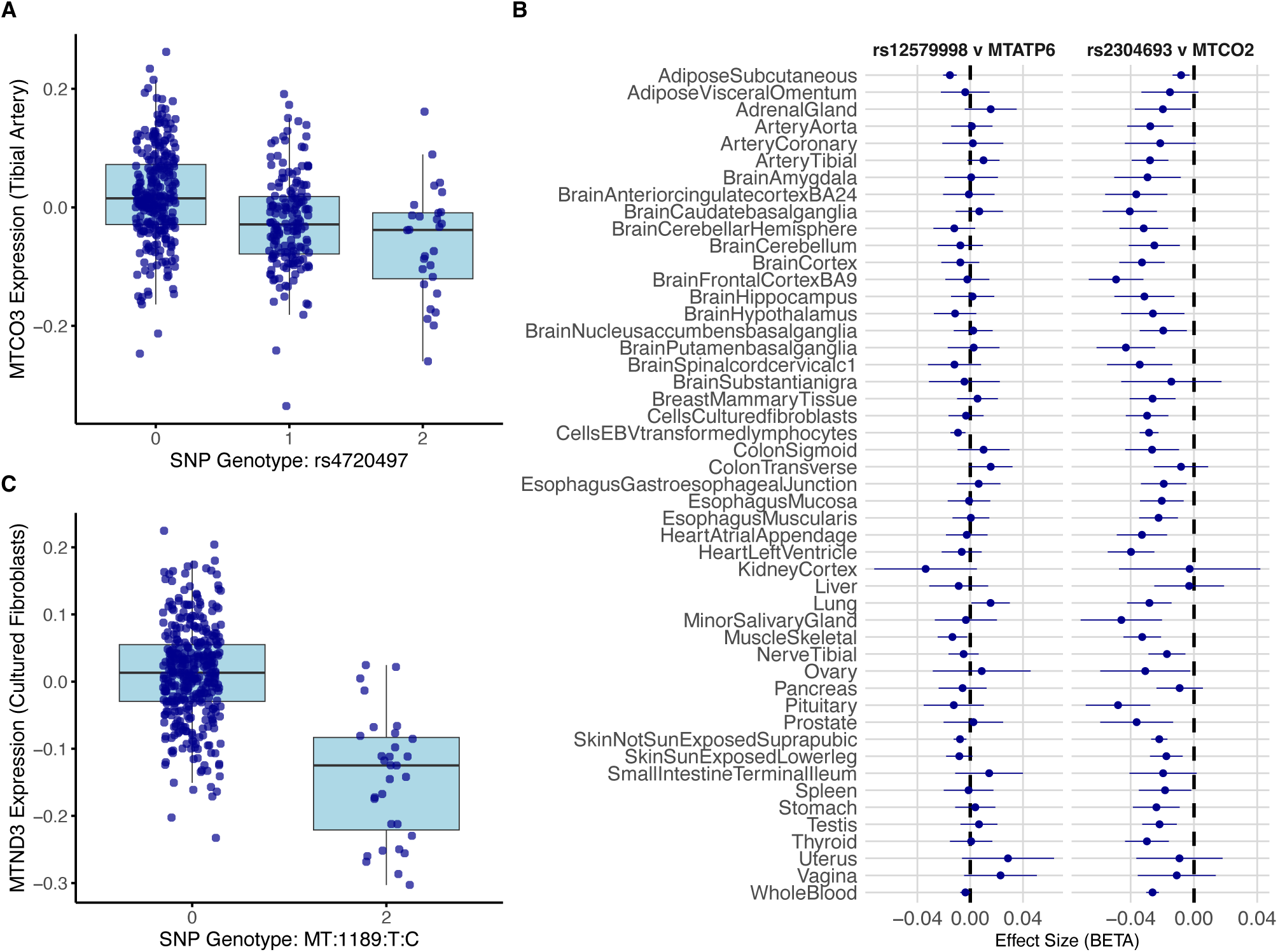
Visualisation of eQTL Data. (A) Example cis-eQTL plot showing the association between a nuclear genetic variant and residual expression of the mitochondrial gene MTCO3 in tibial artery tissue. The variant is located near the nuclear gene FASTKD4, which mediates its eUect; (B) Tissue specificity of nuclear–mitochondrial eQTLs: the left panel shows a tissue-specific association between rs12579998 and MTATP6 expression, with the strongest eUect in subcutaneous adipose tissue; the right panel shows a broadly consistent association between rs2304693 and MTCO2 across multiple tissues, suggesting a shared regulatory mechanism; (C) Example of a mtDNA eQTL: a variant within the mitochondrial gene MTRNR1 is associated with expression of MTND3 in cultured fibroblasts, indicating that polymorphisms in mtDNA can influence mitochondrial gene expression.

**Table 1:** Nuclear genetic variants associated with mtDNA-encoded transcript abundance. For each genetic variant, the nearest nuclear gene is annotated. Where the lead variant is functional or in linkage disequilibrium (LD) with a functional variant, the gene in which the variant resides or exerts its eUect is reported. In cases where the variant is predicted to mediate mtDNA transcript abundance through regulatory influence on a proximal nuclear gene, that gene is also indicated.

| Tissue | Rs ID | Annotation | P value | Gene | Closest Gene | Functional Variant | Mediation |
| --- | --- | --- | --- | --- | --- | --- | --- |
| Adipose | rs12579998 | intergenic | 1.9E-15 | MTND4 | MRPS35 | MRPS35 |  |
| Adipose | rs12579998 | intergenic | 2.4E-27 | MTND1 | MRPS35 | MRPS35 |  |
| Adipose | rs12579998 | intergenic | 1.4E-11 | MTATP6 | MRPS35 | MRPS35 |  |
| LCL | rs2304694 | missense | 4.9E-12 | MTCO2 | FASTKD4 | FASTKD4 |  |
| LCL | rs2304693 | missense | 3.6E-13 | MTND4 | FASTKD4 | FASTKD4 |  |
| Skin | rs2304693 | missense | 6.8E-17 | MTCO2 | FASTKD4 | FASTKD4 |  |
| Skin | rs12833700 | intron | 8.5E-19 | MTND1 | MRPS35 | MRPS35 |  |
| Skin | rs2304693 | missense | 4.5E-22 | MTCO3 | FASTKD4 | FASTKD4 |  |
| Whole Blood | rs12616982 | intron | 2.6E-38 | MTND6 | PNPT1 | CFAP36/PNPT1 |  |
| Whole Blood | rs2304693 | missense | 1.5E-38 | MTCO2 | FASTKD4 | FASTKD4 |  |
| Whole Blood | rs782637 | intron | 2.1E-09 | MTCYB | PNPT1 | PNPT1 |  |
| Whole Blood | rs7090805 | intergenic | 1.5E-10 | MTCYB | MTPAP | NA | None |
| Whole Blood | rs6725101 | intron | 4.4E-46 | MTND5 | PNPT1 | CFAP36/PNPT1 |  |
| Whole Blood | rs41304800 | missense | 5.6E-16 | MTCO1 | FASTKD5 | FASTKD5 |  |
| Whole Blood | rs7600434 | intron | 3.8E-10 | MTND3 | KLHL41 | NA | KLHL41 |
| Whole Blood | rs2304693 | missense | 1.5E-25 | MTND4 | FASTKD4 | FASTKD4 |  |
| Whole Blood | rs2304694 | missense | 1.1E-25 | MTND1 | FASTKD4 | FASTKD4 |  |
| Whole Blood | rs6973982 | intron | 6.1E-15 | MTATP6 | FASTKD4 | FASTKD4 |  |
| Whole Blood | rs782634 | intron | 2.9E-11 | MTCO3 | PNPT1 | PNPT1 |  |
| Whole Blood | rs2304693 | missense | 2.4E-30 | MTCO3 | FASTKD4 | FASTKD4 |  |
| Whole Blood | rs2245761 | intron | 8.3E-10 | MTRNR2 | MTPAP | MTPAP |  |
| Tibial Artery | rs4720497 | upstream | 2.5E-13 | MTCO3 | FASTKD4 | NA | FASTKD4 |
| Putamen | rs75141110 | intron | 1.0E-09 | MTND1 | ALK | NA | None |
| Fibroblasts | rs11366537 | upstream | 8.6E-12 | MTND4 | MYO1G | FASTKD4 |  |
| Skel. Muscle | rs13399762 | intron | 2.9E-09 | MTND5 | PNPT1 | CFAP36/PNPT1 |  |

To assess the gene and tissue specificity of these associations, we first asked whether each of the 25 peak genetic variants was also associated with other mtDNA-encoded genes within the same tissue type. Considering all 350 possible variant–gene combinations (25 variants x 14 other mtDNA genes), we found that 58% showed nominal associations (P<0.05), suggesting a mixture of both broad and gene-specific regulatory effects. For example, rs12579998 (near *MRPS35*) in subcutaneous adipose tissue was associated with the expression of 14 mtDNA-encoded genes, whereas rs13399762 (within *PNPT1*) in skeletal muscle was associated only with *MTND5* and *MTND6*. We next evaluated tissue specificity by testing whether each of the 25 peak genetic variants was associated with the same mtDNA-encoded gene across other GTEx tissue types. Of the 1,200 possible variant and tissue combinations (25 variants x 48 tissues), approximately half showed nominally significant associations. Some variant–gene associations were broadly replicated across tissues. For instance, rs2304693, a missense variant in *FASTKD4*, was associated with *MTCO2* in 39 different tissue types (Figure 1B). In contrast, other associations were highly tissue specific. For example, the association between rs12579998 and *MTATP6* was largely restricted to adipose tissue, with additional associations observed in only four other tissues, one of which showed an opposite direction of effect (Figure 1B). Together, these results suggest that nuclear regulation of mtDNA-encoded transcript abundance involves both globally acting and gene- or tissue-specific mechanisms.

Finally, given that mtDNA-encoded gene expression levels are considered to be highly correlated along mtDNA and across tissues^18,24^, we also apply a less stringent correction method and identify an additional 58 trans-genome eQTLs at FDR 5% (supplementary table 2). Applying the same functional annotation approaches as above, we linked peak nuclear genetic variants to several known mitochondrial RNA processing genes, including *MRPP3*, *LONP1* and *ELAC2*, as well as other genes with unknown links to mitochondria that may warrant further investigation (*MYOG*, *NACAD*, *REP15* and *CCM2*).

### mtDNA genetic control of mtDNA-encoded transcript abundance

Within GTEx data, we also performed association analyses between fixed genetic variants in mtDNA itself (homoplasmies, MAF>5%) and the transcript levels of mtDNA-encoded genes in each tissue type. In total, we found 15 cis-genome eQTLs (peak mtDNA genetic variant-gene expression pairs) for mtDNA-encoded transcript abundance after Bonferroni correction (P<5x10^-8^/15, supplementary table 3). Most of these associations (9 out of 15) occured between a genetic variant in *MTRNR1* (1189T-C) and the expression levels of *MTND3* across different tissue types (Subcutaneous Adipose, Brain Cortex, Cultured Fibroblasts (Figure 1B), Esophagus (Gastroesophageal Junction, Mucosa, and Muscularis), Lung, Nerve Tibial, and Pituitary) reflecting a broad regulatory effect. Notably, the same genetic variant has also been found to be associated with diverse clinical phenotypes, including primary bilateral coxarthrosis (M16.0), ill-defined heart disease (I51.8), and lymphocyte count^10^, suggesting a model in which genetic variation in mtDNA contributes to complex, multi-organ phenotypes through shared pathways of mitochondrial function.

### Genetic prediction of mtDNA-encoded transcript abundance

To build genetic scores that predict mtDNA-encoded transcript abundance, we focused on individuals of European ancestry, who comprise the vast majority of the GTEx dataset. For each of the 15 mtDNA-encoded genes, we constructed multiple prediction models across 49 tissue types using both nuclear and mtDNA genetic variants. Genetic variants were selected based on different P-value thresholds, and we applied a range of machine learning approaches (see Methods). Following model construction, all 735 gene–tissue combinations yielded at least one viable prediction model that included at least one genetic variant.

To test the predictive accuracy of our genetic scores for mtDNA-encoded transcript abundance, we performed two validation exercises. First, we conducted internal validation using GTEx data. For 46 tissue types with at least 100 samples, we split the data into training (80%) and validation (20%) subsets. Genetic scores were built using the training data and then used to predict transcript abundance in the validation set, which was then compared to observed expression values. Of the 690 possible gene-tissue combinations (46 tissues x 15 genes), 687 yielded at least one viable model. The mean R^2^ for the best model per gene-tissue pair was 0.071 (Figure 2A), significantly higher than expected by chance (P < 1x10^-6^), and comparable to recent cis-eQTL-based approaches (e.g., the OmicsPred study^25^, which reported average R^2^ = 0.06 in whole blood). Notably, 147 models had R^2^ > 0.10, and 12 exceeded R^2^ > 0.30 (Figure 2B). *MTND3* had the highest mean R^2^ across tissues (0.089), while *MTND2* had the lowest (0.061).

**Figure 2:**
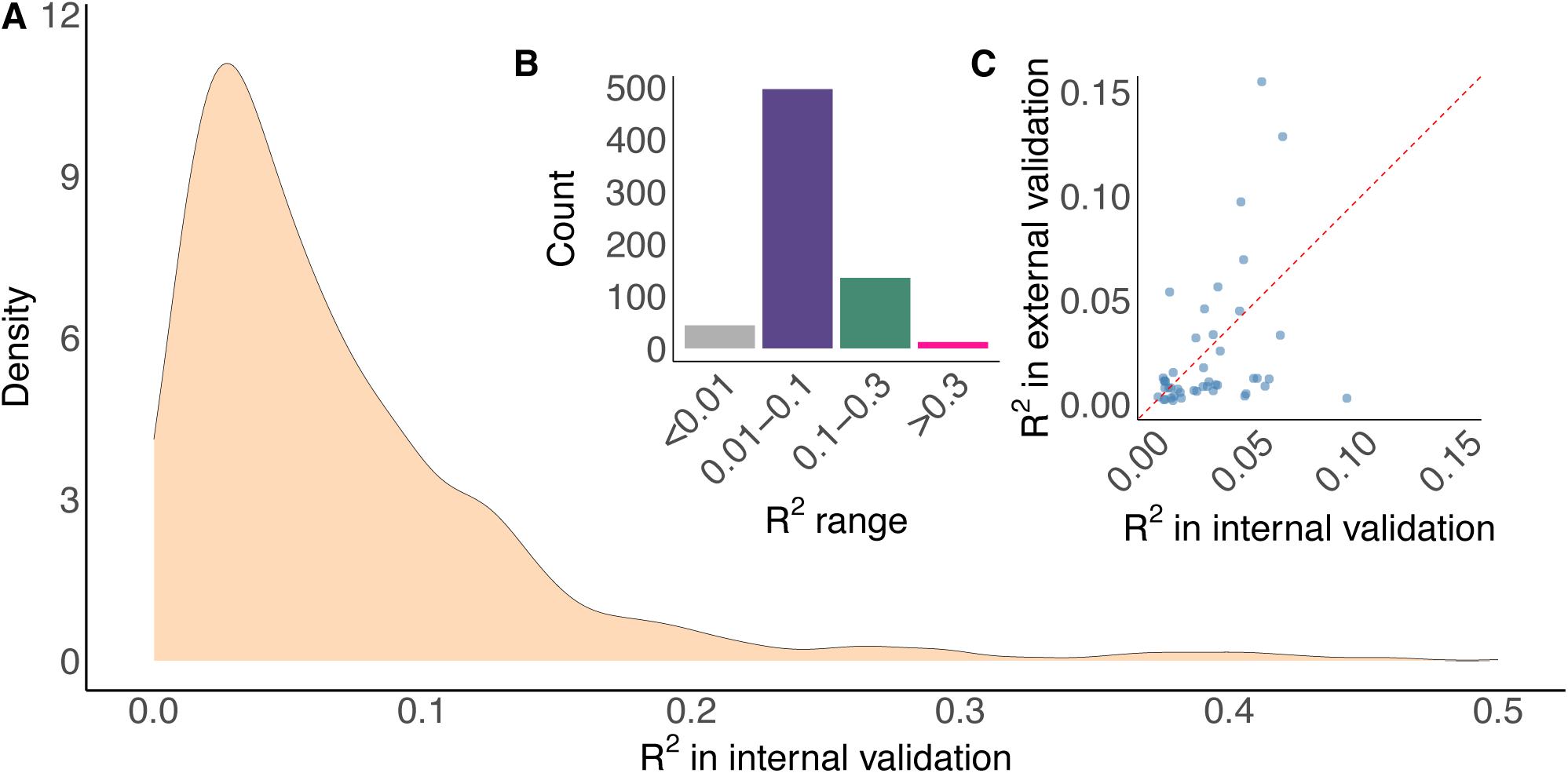
The distribution of R^2^ values from internal validation, where predicted mtDNA-encoded expression values were compared against observed values (A). The number of models within each R^2^ bin for internal validation (B, inset), and the correlation between internal and external R^2^ values for the same genes and tissues (C, inset).

Second, we assessed external validity by applying the full GTEx-trained models to three independent datasets (whole blood, subcutaneous adipose and non-sun exposed skin). Predicted transcript abundances were compared to observed values obtained from RNA sequencing data. The mean R^2^ for the best model per gene-tissue pair was 0.023, again significantly above chance (P < 1x10^-6^) and comparable to previous work predicting nuclear gene expression from local variants (e.g. Gamazon et al. (2015)^26^, R^2^ = 0.0458 in whole blood; 0.0197 in LCLs). Of the 45 tested models, 12 were significant after Bonferroni correction (P<0.05/45), with an additional 13 reaching nominal significance (P<0.05). *MTND3* again showed the highest average R^2^ (0.099) and *MTCYB* the lowest (0.005), and R^2^ values are significantly correlated between internal and external validation tests (P=0.009, Pearson’s R=0.38, Figure 2C). These results demonstrate that mtDNA-encoded gene expression can be reliably predicted from genetic data across multiple tissue types and independent cohorts.

### Phenome-wide Association Analysis with Disease Traits in UK Biobank Data

Next, we applied our genetic scores to predict mtDNA-encoded transcript abundances in unrelated individuals of European ancestry from the UK Biobank (UKBB; N=377,439). We then assessed associations between transcript abundances and the incidence of 699 binary disease traits, each with at least 500 cases in UKBB (supplementary table 4). Specifically, we tested associations between each disease trait and transcript abundance for all 15 mtDNA-encoded genes across 49 tissue types.

After correcting for multiple testing, we identified 48 significant associations (table 2, supplementary table 5). These include well-established mitochondria-related traits such as type 2 diabetes and ptosis of the eyelid, as well as strong associations with disorders of iron metabolism (which aligns with known mitochondrial roles in iron homeostasis). In addition, we uncovered novel associations with conditions such as coeliac disease, breast cancer and hypertension (see below), suggesting broader relevance of mtDNA gene expression processes in common disease risk.

**Table 2:** Diseases in UKBB that are significantly associated with predicted mtDNA-encoded transcript abundance after Bonferroni correction. Relevant tissues and mtDNA-encoded genes are shown, together with the strongest P-value associated with the disease across all tissues, genes and models.

| mtDNA Gene(s) | Trait | P-value | Tissue(s) |
| --- | --- | --- | --- |
| 6 Genes | E83.1: Disorders of iron metabolism | 1.73E-40 | 7 Tissue Types |
| MTRNR2 | Diabetes (diagnosed by doctor and E11.9) | 6.17E-11 | Spleen |
| MTND1 | Hypertension (including I10) | 1.57E-13 | Aorta (artery) |
| 4 Genes | K90.0: Coeliac disease | 2.15E-11 | 4 Tissue Types |
| MTND1 | H02.4: Ptosis of eyelid | 8.31E-09 | Thyroid |
| MTND4 | C50.4: Malignant neoplasm (Upper-outer quadrant of breast) | 6.88E-14 | Skin (Sun Exposed) |
| MTND4 | C50.9: Malignant neoplasm (Breast, unspecified) | 5.49E-51 | Skin (Sun Exposed) |
| MTND4 | D05.1: Intraductal carcinoma in situ | 1.59E-09 | Skin (Sun Exposed) |
| MTND4 | C77.3: Secondary and unspecified malignant neoplasm: Axillary and upper limb lymph nodes | 9.66E-19 | Skin (Sun Exposed) |
| MTND4L | K57.3: Diverticular disease of large intestine without perforation or abscess | 9.88E-13 | Brain (Spinal cord cervical c1) |
| MTND4L | I83.9: Varicose veins of lower extremities without ulcer or inflammation | 6.57E-09 | Brain (Putamen, basal ganglia) |

#### Associations with hypertension

Predicted *MTND1* transcript levels in aortic artery tissue are significantly associated with hypertension (P = 1.57 x 10^-13^). Although mitochondrial dysfunction has been proposed to contribute to oxidative stress, renal damage and cardiovascular disease^27^, previous studies have reported inconsistent associations between mtDNA genetic variation and blood pressure traits, with recent large-scale analyses finding no significant associations^10,28^. Our findings therefore suggest an alternative mechanism by which *MTND1* expression, rather than mtDNA sequence variation, may influence hypertension risk.

Our genetic score model for *MTND1* transcript abundance was determined by six genetic variants. Notably, one of these variants, rs751891, was strongly associated with hypertension (P = 8 x 10^-56^), and in linkage disequilibrium (R^2^ = 0.6) with the lead variant discovered in a large hypertension and cardiovascular disease genome-wide association study^29^ (GWAS, rs72831343; P = 1.4 x 10^-67^). When rs751891 was excluded from the genetic model, the association between predicted *MTND1* expression and hypertension was no longer significant (P = 0.78), indicating that the observed association is likely driven by this single variant. Further supporting this, rs751891 is an eQTL for *MTND1* specifically in aortic artery tissue in GTEx (P = 9.4 x 10^-6^), and not in any other tissue after correcting for multiple testing (P > 0.05/49). Neither rs751891 nor the linked rs72831343 were eQTLs or splice QTLs (sQTLs) for nuclear-encoded genes in GTEx v8 (European subset, aortic artery), although rs72831343 was an eQTL for the gene *CABCOCO1* in the full GTEx v10 dataset (P = 6.6 × 10^-5^), which includes individuals of all ancestries.

To further investigate causality, we conducted a GWAS for hypertension in unrelated European individuals from the UK Biobank and performed colocalization analysis. We found evidence of colocalization between the *MTND1* expression signal in aortic artery and hypertension (posterior probability of shared signal, h_4_ = 0.75, Figure 3A). In contrast, none of the nuclear genes within 1MB of rs751891 showed colocalization (all h_4_ < 0.12; *CABCOCO1* h_4_ = 0.0002), suggesting that nuclear genetic variants associated with hypertension may mediate their effect by regulating *MTND1* expression in the vascular wall.

**Figure 3:**
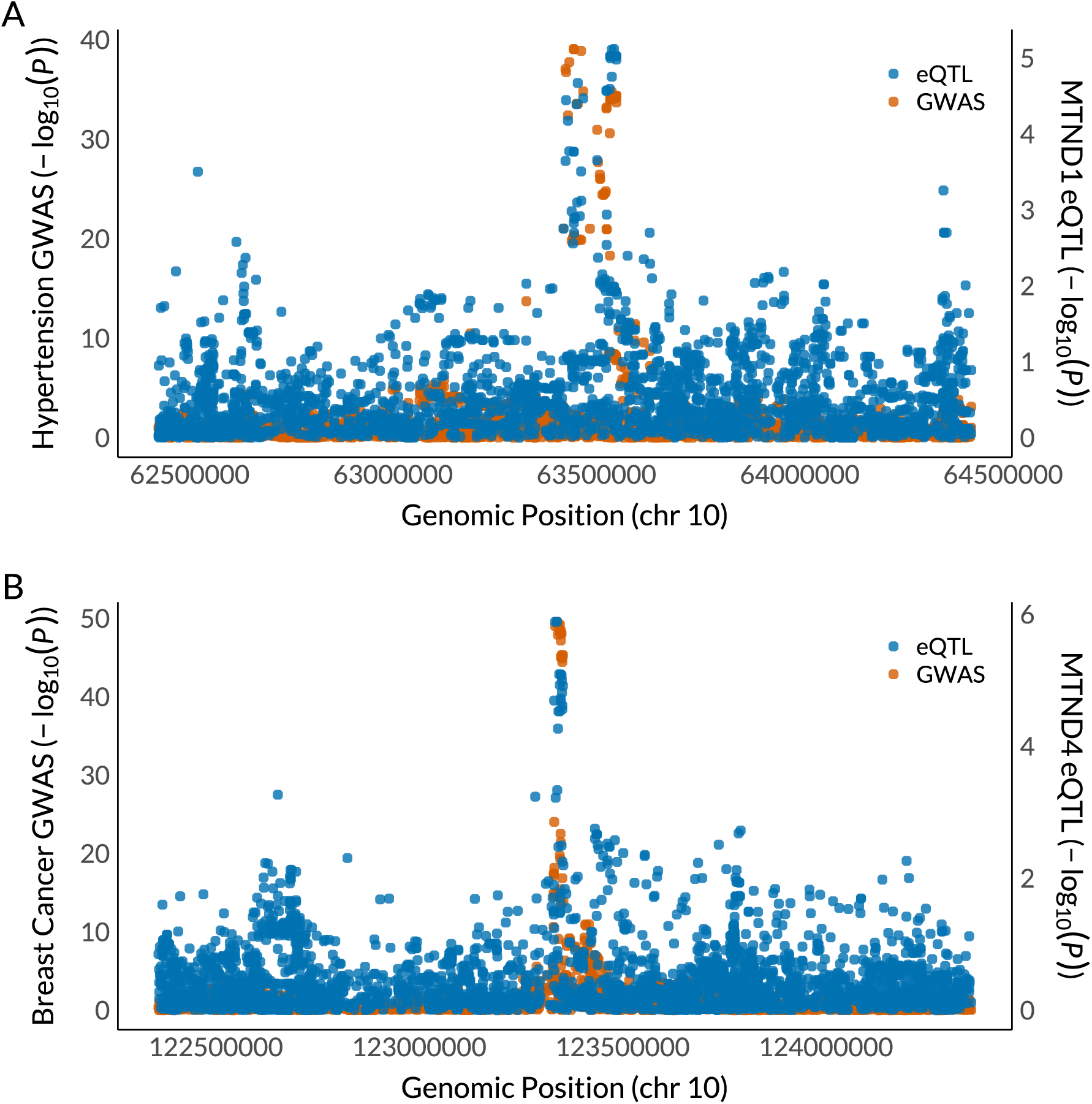
Colocalization plots between (A) expression of MTND1 in the Aorta Artery and hypertension in UK Biobank (UKBB), and (B) expression of MTND4 in sun-exposed skin and malignant neoplasm of the breast (breast cancer) in UKBB.

#### Associations with Breast Cancer

Predicted expression levels of *MTND4* in skin (sun-exposed) were strongly associated with several breast neoplasm phenotypes, with the most significant association observed for C50.9 (Malignant neoplasm of the breast, unspecified; P = 5.49 x 10^-51^). While this association arises in skin tissue, it may reflect shared transcriptional signatures across epithelial lineages. Although we find associations between predicted *MTND4* expression and C50.9 in breast tissue at nominal significance, none of these associations pass multiple testing correction (lowest P=0.03), potentially reflecting the smaller number of breast tissue samples in GTEx (n=329) compared to skin (n=508), and the heterogeneity of breast tissue samples which may be dominated by adipocytes rather than epithelial cells, thus diluting expression signals in bulk tissue.

The best-performing genetic score model in skin included five genetic variants. The variant with the largest weight in the model was rs1078806, located within the *FGFR2* locus, a well-known susceptibility region for breast cancer^30^. Removal of this variant from the model abolished the association between predicted *MTND4* expression and C50.9 (P = 0.94), suggesting that this variant is primarily driving the observed signal. This lead genetic variant was significantly associated with *MTND4* expression in skin (sun-exposed) in GTEx (P = 1.26 x 10^-6^) but is not associated with *MTND4* expression in any other tissue after correction for multiple testing (P > 0.05/49). rs1078806 is not annotated as an eQTL or sQTL for any nuclear gene in skin or breast tissue in either GTEx v8 (European subset) or GTEx v10 (full dataset), including *FGFR2*.

To investigate potential causality in our data, we assessed colocalization between the *MTND4* signal in skin and C50.9 GWAS results from unrelated European individuals in UK Biobank. This revealed strong colocalization for *MTND4* (h_4_ = 0.99) but not for *FGFR2* (h_4_ = 0.007) or any other nuclear gene within 1MB of the peak variant, reinforcing the potential relevance of *MTND4* regulation in disease risk.

### Phenome-wide Association Analysis with Quantitative Traits in UK Biobank Data

Next, we assessed associations between mtDNA-encoded transcript abundance derived from genetic scores, and 127 health-related quantitative traits in UK Biobank, encompassing anthropometric measures, blood cell indices, and a range of serum and urinary biomarkers (supplementary table 6). In total, we identified 1,845 significant associations (after multiple-testing correction, supplementary table 7). Notably, several traits showed signals consistent with previous findings from direct association testing of mtDNA genetic variants^10^, particularly those related to kidney function (e.g. creatinine, cystatin C, eGFR, urea), liver enzymes (e.g. AST, ALT), a broad array of haematological traits (e.g. red and white blood cell counts, platelet traits), airway function and height, reinforcing the role of mitochondrial variation in these physiological domains. Beyond these, our expression-based approach revealed associations with a much wider spectrum of traits, including metabolic biomarkers (e.g. HDL cholesterol, triglycerides, glucose), reproductive and endocrine measures (e.g. testosterone levels, age at menopause, reproductive span), inflammatory markers (e.g. C-reactive protein), blood pressure, a wider range of anthropomorphic traits (e.g. waist-to-hip ratio, weight) and indicators of physical performance (e.g. grip strength).

Overall, these associations exhibited a range of tissue specificity (Figure 4). Certain traits, such as bilirubin levels and height, were associated with mtDNA transcript abundance across many different tissues, potentially reflecting widespread mitochondrial involvement or the systemic nature of these phenotypes. In contrast, other traits showed marked tissue specificity. For instance, associations between glucose levels and mtDNA transcript abundance were observed only for the pancreas, and testosterone levels were linked to transcript abundance only in liver (the hormone’s primary site of metabolism), both of which are consistent with known physiological functions. In other cases, the relevance was less clear. Associations involving albumin and phosphate, for example, were identified only in brain regions, where their functional significance is not well understood. Together, these patterns suggest that organ-specific traits may be shaped by tissue-specific regulation of mtDNA-encoded transcripts, likely mediated by nuclear factors involved in mitochondrial RNA processing.

**Figure 4:**
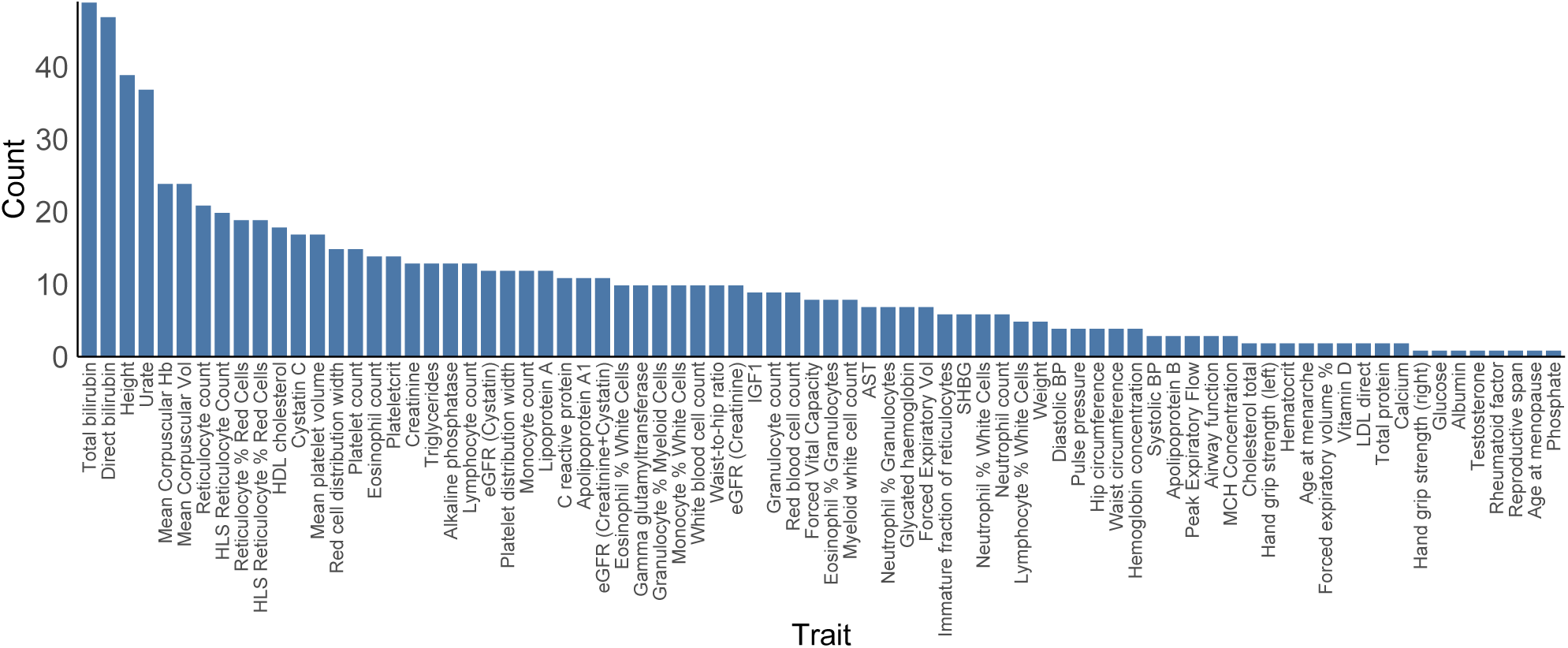
Number of tissues with significant associations between genetically predicted mtDNA transcript abundance and each trait.

#### Liver, Kidney and Lung function

Among liver-related biomarkers, total and direct bilibrubin levels were most strongly associated with mtDNA-encoded transcript levels derived from genetic scores (most significant association across genes and tissues, P=5.4x10^-302^), particularly those involving *MTATP8* (four of the top 10 most significant associations with the trait). Bilirubin is a byproduct of haem catabolism and is processed in mitochondria-rich hepatocytes which are heavily dependent upon ATP synthesis, making its sensitivity to mitochondrial gene regulation biologically relevant. We also observed associations with AST and GGT, which are both sensitive markers of hepatocyte injury, raising the possibility that that variable mtDNA transcriptional processes contribute to underlying hepatocellular stress. For kidney-related biomarkers, the strongest association was between *MTND3* transcript abundance in the liver and urate levels (P=5.7x10^- 136^). While urate is excreted renally, it is produced through xanthine metabolism in the liver and other tissues, implicating mitochondrial control over purine metabolism, which is critical for DNA synthesis. Cystatin C and eGFR (Cystatin-based) were also strongly associated with several mtDNA genes, the most significant association being with *MTRNR1* in subcutaneous adipose tissue (P=9.7x10^-101^), suggesting systemic mitochondrial influences on filtration markers and indicating crosstalk between energy-storing tissues and renal clearance mechanisms. Associations with Creatinine and Phosphate were present but weaker, aligning with the idea that not all renal biomarkers are equally sensitive to variation in mitochondrial function. In terms of lung function, mtDNA transcriptional processes were most significantly associated with forced vital capacity (FVC) and forced expiratory volume in 1 second (FEV1), particularly for genes such as *MTCO2* (lowest P=5.0x10^-25^). These traits are influenced by airway structure, pulmonary elasticity and systemic inflammation, all of which can be impacted by mitochondrial bioenergetics and oxidative stress pathways.

#### Cardiovascular and metabolic function

Traits linked to cardiovascular function, particularly lipids, yielded some of the strongest and most reproducible associations with mtDNA-encoded genes. Among these, *MTND2* had highly significant associations across HDL cholesterol, Apolipoprotein A1, LDL, and Triglycerides (lowest P=3.2x10^-103^). These findings align with known mitochondrial involvement in β-oxidation, lipoprotein remodelling and cholesterol efflux, emphasising the role of mitochondria in lipid homeostasis. Furthermore, transcript abundances of several mtDNA-encoded genes were associated with C reactive protein levels (lowest P=7.2x10^-70^), which is associated with inflammation and atherosclerosis. Systolic blood pressure (SBP), diastolic blood pressure (DBP) and pulse pressure showed associations with several mtDNA genes (lowest P=3.7x10^-12^), including *MTND1* in the Aorta artery, independently endorsing the association with hypertension described above. Finally, glucose and HbA1c levels showed significant associations with multiple mtDNA-encoded genes, most notably *MTCO1* (lowest P=1.4x10^-12^). These genes likely influence glucose homeostasis via mitochondrial roles in insulin secretion, oxidative phosphorylation and redox buffering. The influence of mitochondrial function on glycaemic biomarkers reinforces the hypothesis that mitochondrial dysfunction plays a role in the pathophysiology of type 2 diabetes. This is further supported by evidence linking mtDNA variation to glycaemic and lipid-related traits^31^.

#### Blood cell traits

mtDNA-encoded genes showed broad and highly significant associations across multiple blood cell indices, suggesting that haematopoiesis and blood cell homeostasis are deeply influenced by mitochondrial function. Among the most consistent signals across genes and tissues were associations with mean corpuscular hemoglobin (MCH) and mean corpuscular volume (MCV) (lowest P=2.4x10^-25^). These findings are biologically coherent, as mitochondrial function is critical during erythropoiesis for haem biosynthesis, iron utilization and redox regulation. White cell parameters, including white blood cell (WBC) count, neutrophil count, monocyte count, and lymphocyte count, were associated with several mtDNA-encoded genes (lowest P=8.6x10^-40^). These associations may reflect the role of mitochondria in immune cell activation, differentiation and lifespan, as well as immunometabolism programming, especially relevant in lymphocytes and monocytes where mitochondrial metabolism regulates cytokine production and memory cell function.

#### Anthropomorphic and age-related traits

Finally, mtDNA-encoded transcript abundances showed robust associations with multiple anthropometric traits, particularly height and weight, suggesting that mitochondrial function plays a key role in growth, body composition and energy regulation. Among the strongest associations was the link between *MTND2* in visceral adipose and height (P=3.7x10^-28^). Related traits such as waist-to-hip ratio were also associated with multiple mitochondrial genes, notably *MTND3*. Associations with grip strength (a well-established biomarker of biological aging, muscle function and frailty risk) were also notable for both left- and right-hand grip strength.

### Associations with age-related common diseases

Given the limited number of late-onset disease cases in the UK Biobank, we conducted a summary statistic-based transcriptome-wide association study (TWAS) to evaluate whether transcript levels derived from genetic scores of the 15 mitochondrial DNA-encoded genes across 49 GTEx tissues were associated with risk for five common age-related diseases: type 2 diabetes (T2D), Parkinson’s disease (PD), atrial fibrillation (AF), coronary artery disease (CAD), and Alzheimer’s disease (AD). Although mitochondrial dysfunction has been implicated in these diseases, prior studies have reported that nuclear-encoded mitochondrial genes were not strongly enriched for common variant heritability in GWAS^32^. Here, we sought to revisit this question using mtDNA transcript levels as a molecular intermediate that could reveal regulatory contributions to disease risk.

We observed the most robust associations for T2D, with twelve transcriptome-wide significant gene–tissue associations (Bonferroni-corrected P<5x10^-6^) involving eight mitochondrial genes. The strongest signal was for *MTCO2* expression in cultured fibroblasts (Z=−8.92, P=4.7x10^-19^), however we also saw associations for *MTND4* in the transverse colon and *MTCYB* in skeletal muscle, as well as in peripheral tissues like oesophagus and skin. Additional signals were observed in central nervous system tissues, including *MTCO2*, *MTCO3*, *MTND2* and *MTATP6* in the caudate, frontal cortex and nucleus accumbens, although the reasons for these associations are not clear.

For PD, four transcriptome-wide significant associations were detected, including *MTND6* and *MTND1* in the adrenal gland, *MTND4* in coronary artery, and *MTATP8* in the pituitary, suggesting a possible contribution of both endocrine and vascular tissues to disease risk. Notably, the adrenal gland originates from the neural crest and is functionally linked to dopaminergic systems, which are central to PD pathophysiology. Furthermore, the majority of associations involve Complex I genes (MTND1, MTND4, MTND6), echoing longstanding evidence that Complex I dysfunction plays a central role in PD pathogenesis^33^. For AF, three gene-tissue associations reached significance. These included *MTATP8* expression in the spinal cord (cervical C-1) (Z = 5.35, P = 8.8x10^-8^), *MTND2* in liver, and *MTND1* in skeletal muscle (Z = −5.45, P = 5.1x10^-8^), pointing to a role for both central autonomic pathways and peripheral energy metabolism in arrhythmia susceptibility. No significant associations were observed for CAD or AD after multiple testing correction.

## Discussion

In this study, we demonstrate that genetically predicted levels of mtDNA-encoded transcripts are associated with a broad spectrum of human diseases and traits, especially those related to metabolism, cardiovascular function and neurodegeneration. Because our transcript predictions are based entirely on inherited germline variants, the observed associations must reflect the influence of genetics (and by extension, mtDNA transcriptional processes) on disease risk, and not the reverse. This genetic anchoring provides a key advantage over direct measurement of transcript levels, which are often confounded by environmental, temporal or disease-related effects.

In total, we identified 25 significant trans-nuclear genetic associations with mtDNA-encoded gene expression, along with numerous additional signals at a 5% FDR. Many of the peak genetic variants map to nuclear genes with well-established roles in mitochondrial RNA metabolism, including *FASTKD4, FASTKD5, PNPT1, MRPP3, ELAC2 and LONP1*. These associations point to RNA processing, rather than transcription initiation alone, as a major determinant of mtRNA abundance. For instance, variants near *MRPP3*, a catalytic subunit of the mitochondrial RNase P complex, were associated with transcript levels of multiple mtDNA genes, consistent with its role in cleaving precursor transcripts. Similarly, variants at the *FASTKD4* locus, known to influence transcript stability and maturation, were linked to several mtDNA-encoded genes. Similarly, while mammalian mitochondrial transcripts are co-regulated through polycistronic transcription, our genetic data suggest that individual mtDNA genes may nonetheless be subject to distinct regulation at the post-transcriptional level. Together, these findings provide strong genetic evidence that post-transcriptional regulation is a central axis of mitochondrial gene expression control in human tissues.

Our results build on previous work showing associations between mtDNA variants and several complex traits^9,10^, but expand the scope by identifying new links, especially with traits relevant to cardiovascular health, metabolism, immune function and aging. Notably, we also observe associations between mtDNA-encoded transcript abundance and age-related diseases such as type 2 diabetes and Parkinson’s disease, despite earlier studies reporting no enrichment of nuclear-encoded mitochondrial gene variation in these conditions^32^. These findings may be particularly relevant given recent studies showing that pharmacological modulation of mtDNA transcription can influence physiological outcomes in mice^34,35^. For example, we observe that reduced transcript levels of specific mtDNA genes in liver are associated with lower waist-to-hip ratio, potentially paralleling results in mouse models where transcriptional inhibition of mtDNA attenuates obesity, hepatosteatosis and restores normal glucose tolerance^34^. This represents the first genetic evidence that such interventions could be effective in humans.

While some associations may reflect differences in statistical power across tissues, others reveal biologically meaningful tissue-specific effects. For instance, *MTND1* expression in aortic artery shows both strong genetic regulation and colocalised association with hypertension, suggesting that local vascular mitochondrial function contributes to blood pressure control. Likewise, *MTCO1* expression in skeletal muscle is genetically linked to type 2 diabetes risk, aligning with the critical role of muscle oxidative metabolism in glucose homeostasis. These findings support the hypothesis that nuclear regulation of mtDNA-encoded transcript abundance contributes to the selective vulnerability of specific tissues to mitochondrial dysfunction, providing a possible explanation for why mitochondrial pathology manifests so differently across organs in both rare mitochondrial syndromes and common age-related diseases.

Several important caveats should be considered. First, RNA sequencing data captures steady-state RNA levels, which reflect not only transcription but also other processes such as mtDNA copy number, RNA processing, modification and degradation. While this limits interpretation of transcriptional activity *per se*, these RNA levels remain biologically meaningful as proxies for mitochondrial output. Second, although we model gene expression separately for each of the 15 mtDNA-encoded genes, mitochondrial transcription operates via polycistronic mechanisms. Some genes (e.g., *MTATP6*/*8*) are bicistronic, and *MTND6*, transcribed from the light strand, is not polyadenylated and thus should be underrepresented in GTEx poly-A+ data. Similarly, mitochondrial rRNAs are minimally adenylated and typically excluded from standard RNA-seq. Nevertheless, we find robust signals for several of these genes, which may reflect alternative processing (e.g., degradation fragments or longer precursor transcripts) and still carry informative biological signal.

Third, while our use of germline genetics helps infer the direction of effect (from gene expression to trait), we cannot fully exclude pleiotropic effects, where model variants influence another pathway that secondarily affects mitochondrial function. This does not undermine the utility of our findings but highlights the need for cautious interpretation, especially when evaluating causality. Finally, in some cases, the tissues showing the strongest association with a trait may not be the most biologically intuitive. This could reflect shared regulatory variants across tissues, differences in sample size and power or unknown aspects of tissue biology. As population-scale single-cell datasets become available, future studies could better resolve these signals at the level of cell type–specific mitochondrial function.

Despite these limitations, our analysis shows that an inherited predisposition to differences in mtDNA transcript abundance contributes to the origin of a broad spectrum of common human diseases and traits. Within this, nuclear modulation of mitochondrial RNA processing plays a critical role, providing compelling novel therapeutic targets for disease treatment or prevention.

## Methods

### Processing of RNA sequencing data

Aligned RNA sequencing data from 15,201 samples were obtained from the GTEx project via dbGaP (phs000424.v8.p2) for 49 different cell/tissue types that had at least 70 samples available with paired genetic data. Aligned data were sorted and converted back to fastq format with samtools^36^ (v1.14), before being realigned to the human reference sequence (GRCh38, primary assembly) with STAR^37^ (v2.7.6) to ensure the consistency of alignments across datasets and to focus on mitochondria-specific mappings. Alignment was performed within STAR using two-pass mode incorporating Gencode gene annotations (v42) and the following parameters: - -outSAMstrandField intronMotif; --outFilterType BySJout; --alignSJoverhangMin 8; -- alignSJDBoverhangMin 1; --outFilterMismatchNmax 999; --outFilterMismatchNoverReadLmax 0.05; --alignIntronMin 20; --alignIntronMax 1000000; --alignMatesGapMax 1000000. The ‘-- quantMode GeneCounts’ flag was used to obtain gene counts for all genes, focussing on properly paired and uniquely mapped reads only, thus minimizing the likelihood of including incorrectly placed reads (particularly those associated with NUMT sequences) in gene count data. After alignment, samples that had an intergenic mapping rate >30%, an overall base mismatch rate >1%, a ribosomal RNA mapping rate >30%, total reads < 5,000,000 or had zero reads for any ribosomal or protein coding gene encoded within mitochondrial DNA (mtDNA) were removed using in house scripts and RNAseQC^38^ (v2.4.2). Finally, raw gene counts were converted to transcripts per million (TPM) and log_10_ transformed. Distributions of all genes with mean TPM>2 per sample were plotted and visual outlier samples were removed. Principal components using the same data were also calculated and visual outlier samples were removed. After quality control steps, this left a total of 15,066 samples for analysis.

### Processing of genetic data

Variant call format (VCF) files generated from whole genome sequencing of 838 individuals from the GTEx project were obtained via dbGaP (phs000424.v8.p2). Variants were filtered initially for variant and genotype quality >40, minor allele frequency >1%, a maximum missing rate of 1% and keeping only those in Hardy-Weinberg equilibrium (HWE) (P>0.001). As VCF files did not include genetic variants encoded in mtDNA, CRAM files containing whole genome sequencing data were downloaded and reads that aligned to mtDNA were converted to BAM and then FASTQ format with SAMtools^36^ (v1.10). Raw reads were then trimmed with Trim Galore (v0.4.0) (stringency 3, quality 20), before being aligned to the human reference sequence (GRCh38, primary assembly) with BWA-MEM^39^ (v0.7.17). Alignment duplicates (marked with Picard tools), non-uniquely mapped or properly paired reads and reads with a mismatch rate greater than 4% were removed. Genetic variants were then called with Mutserve^40^ (v2.0.0-rc13) (using flags --level 0.05 --baseQ 30) and filtered at minor allele frequency (MAF)>1%. Using custom software, genetic variants were then removed in regions that are known to be difficult to align to (66-71bp, 300-316, 513-525, 3106-3109, 12418-12425, 16182-16194), and heteroplasmies that are known to be common in the population were removed. Finally, heteroplasmies where the non-reference allele was present at frequency >95% were converted to non-reference homplasmies, and those where the non-reference allele was present at frequency <5% were converted to reference allele calls, before mtDNA genetic variation data were merged with nuclear genetic data.

### Quantitative trait loci mapping

Expression quantitative trait loci (eQTL) mapping was performed separately for each of the 15 genes encoded in mtDNA (13 protein coding and 2 rRNA), in each of the 49 tissue/cell types. For each tissue/cell type, TPMs for all genes were extracted (removing genes with zero expression in any individual), and then log_10_ transformed and median normalised. PEER^22^ factors were calculated per tissue dataset using all genes that had mean TPM>2 across samples. Genetic principal components (gPC) were calculated using corresponding genetic data, using only individuals that had expression data available for each given tissue. Nuclear genetic variants with MAF>5% and a maximum missing rate of 1% were selected, and genetic variants in high LD were removed with PLINK (v1.9) (--indep-pairwise 100 5 0.3), before running smartpca in Eigensoft^41^ (v8.0.0) to calculate 10 gPCs for each sample. Expression values for genes encoded in mtDNA were then extracted, masking outliers that were further than 3x the inter-quartile range (IQR) outside the central 50% of the data.

Association analysis was performed for each gene separately using a linear model within PLINK, controlling for 10 PEER factors and 5 gPCs. For four tissue types, we have previously performed QTL mapping in other independent datasets^18^, including data from TwinsUK^42^ (whole blood, n=363; subcutaneous adipose, n=652; non-sun exposed skin, n=651; and lymphoblastic cell lines (LCLs), n=733), the Geuvadis project^43^ (LCLs, n=435), CARTaGENE^44,45^ (whole blood, n=799) and the NIMH resource^46^ (whole blood, n=903). For these tissue types, we performed meta-analysis in PLINK using a fixed effects model combining newly derived association statistics from GTEx data as described above, and summary statistics from analysis described in Ali *et al* (2019)^18^. For each tissue, false discovery correction (Benjamini-Hochberg) was applied to raw p-values within each dataset by merging all 15 genes and genetic variants, following the approach applied by the GTEx consortium^21^. We also selected significant associations based on Bonferroni correction, taking a standard genome-wide significance threshold of 5x10^-8^ and then dividing by 15 (P<3.3x10^-9^). In all association analyses we defined the peak association as the genetic variant with the lowest p-value within a block of 1 MB.

### mtDNA-encoded gene expression prediction

To build expression prediction models for mtDNA-encoded genes we utilised the same data as was used to identify eQTLs, focussing only on individuals of European ancestry as defined by a clustering of samples on the first and second axes of genetic principal components. For each gene in each tissue, we built nine prediction models by selecting nuclear and mitochondrial genetic variants that were associated with the expression of the corresponding gene at three P-value thresholds (all variants, P<0.01, P<1x10^-5^) using three different machine learning approaches (LASSO, Elastic Net and BLUP), as implemented in FUSION^47^. For each prediction model, we first selected nuclear genetic variants on the LD reference panel supplied by FUSION (HapMap3 variants), before filtering genetic variants for P-values. For mtDNA genetic variants, we clumped data using PLINK (with r^2^ of 0.8) based on P-values from eQTL association data, thresholding at the appropriate P-value level for the given model. We then extracted mtDNA expression levels for the given gene and tissue, regressed them against the same covariates used in eQTL analysis, and then used residuals together with corresponding genetic data within FUSION (using --models enet,blup,lasso and --hsq_p 1). For models with P-value thresholds of 0.01 or 1, we also used --hsq_set 1, since the large numbers of genetic variants used in these models were often predicted to have a heritability greater than 1.

### Model validations

To test our ability to predict mtDNA-encoded expression levels from common genetic variation, we performed two types of validation. First, for internal validation we split samples from tissue types with at least 100 samples (46 tissue types) into training and testing data (80 and 20% of individuals respectively). For training data, we recalled eQTLs using genetic and expression information from these individuals for 15 genes encoded in mtDNA, following the methods described above for the full dataset. We then built mtDNA expression prediction models for each gene, selecting genetic variants at three P-value thresholds (all variants, P<0.01, P<1x10^- 5^) and using three different machine learning approaches (LASSO, Elastic Net and BLUP) as described above. After this, for the 20% testing data, we imputed mtDNA expression values using each prediction model and then compared these to observed expression values from RNA sequencing data after regressing out covariates as above. For each gene and tissue, we calculated the Pearson’s correlation coefficient between predicted and observed data and then selected the best R^2^ value across the nine models per gene. To test whether this R^2^ distribution was significantly higher than expected by chance, we performed bootstrapping. For each gene and tissue, we randomly shuffled participant labels on imputed values for each of the nine models, recalculated R^2^ values, and selected best performing model. We combined R^2^ values across all genes and tissues, calculated the mean of this distribution and then tested if this was higher than observed mean R^2^ across all genes and tissues. We repeated this one million times to generate a bootstrapped P value of number of times the mean shuffled R^2^ was higher than mean observed R^2^.

For external validation, we obtained RNA sequencing and corresponding genetic data from the CARTaGENE (whole blood) and TwinsUK (non-sun exposed skin and subcutaneous adipose) cohorts, and processed these data as described in Ali *et al* (2019)^18^. Individuals of European ancestry were defined by a clustering of samples on the first and second axes of genetic principal components. For genetic variants in mtDNA we used RNA sequencing data. Whilst this may not be reliable for heteroplasmies, we reasoned that these data would be robust for detecting fixed genetic variants (homoplasmies). Raw RNA sequencing data were aligned to a reference genome with STAR as described above. Genetic variation in mtDNA was identified using Mutserve (requiring at least 30X coverage), with problem regions removed and heteroplasmies with alternative allele frequency >95% were converted to homoplasmy calls, as above. Genetic variation in mtDNA was then filtered at MAF>1% and a maximum missing rate of 5% before being merged with nuclear data. After filtering and quality control, we used 753 CARTaGENE samples and 401 subcutaneous adipose and 394 non-sun exposed skin samples from TwinsUK (unrelated). Finally for each tissue, we imputed mtDNA-encoded expression values using full prediction models and then compared these to observed expression values from external datasets using a Pearson’s correlation coefficient, after regressing out covariates (5 gPCs and 10 PEER factors). Again, we did this for each of the 9 models per gene, selecting the best performing model in each case, and then used bootstrapping as above to test observed R^2^ values were significantly higher than expected by chance.

### UK Biobank Data

We obtained imputed genotype data from the UK Biobank and converted BGEN files to PLINK binary format (BED) for each autosome. During this conversion, variants were filtered to retain only those with a MAF>1%, call rate >95%, and HWE P-value >0.001. Mitochondrial genotyping data were obtained separately and filtered for MAF >1% and a maximum missing rate of 5%. Genotype datasets were merged across nuclear and mitochondrial genomes using overlapping individuals. SNPs flagged as problematic by PLINK were removed, and individuals who had withdrawn consent (as indicated by negative IDs) were excluded. This resulted in a final analysis set of 488,001 individuals and 4,984,255 genetic variants.

To impute expression values for each individual, we used custom scripts for each of the 9 prediction models (3 methods x 3 P-value thresholds) across all combinations of the 15 mtDNA-encoded genes and 49 tissue types. Each model contained a set of weighted SNPs. Using lifted-over GTEx-based models to match UKBB coordinates (hg19), we calculated imputed expression as the weighted sum of alternate alleles carried by each individual, summing across all SNPs in the model.

Phenotype data were extracted at the same time. We included 169 quantitative health-related traits (see supplementary table 6) and 767 binary disease phenotypes based on self-reported data (field #20002) and ICD10 codes (field #41270), following definitions from Yonova-Doing *et al*. (2021)^10^. Individuals were restricted to those of European ancestry and unrelated, as defined by Pan-UKBB analysis^48^ (Return 2442), and with matching reported and genetic sex. Binary disease traits were further filtered to retain only those with ≥500 cases in this subset, resulting in 699 traits (supplementary table 4). For quantitative traits, data were normalized and filtered according to the parameters described in Yonova-Doing *et al*. (2021)^10^ and described in supplementary table 6.

Haematological traits (n=33) were processed following Astle *et al.* (2016)^49^. This included 15 directly measured traits from Beckman Coulter LH700 Series instruments and 18 derived indices. Blood samples measured >36 hours after venipuncture and samples in the top 4% of mean platelet volume were excluded. Generalized additive models were used to adjust for technical artefacts, including instrument-specific drift and time-of-day, day-of-week, and seasonal effects (modelled with cyclic smoothing). Blood indices were recomputed from adjusted values. Individuals with blood disorders or haematological malignancies were excluded. Biological/environmental covariates (age, sex, BMI, alcohol/smoking history) were adjusted for, stratifying by sex and menopausal status where relevant. Missing covariate values were mean-imputed with dummy indicators. Extreme adjusted values were excluded based on the distribution of raw–adjusted differences and values >4.5 median absolute deviations (MAD) from the median. A PCA was run within each trait group to identify multivariate outliers. Final blood indices were quantile-inverse-normal transformed within hematology analyzer/sex subgroups.

After data preprocessing, we tested for associations between imputed expression levels (per gene x tissue x model) and each phenotype using a linear model. For binary traits, we controlled for age, sex, genotyping batch, and 10 gPCs. For quantitative traits, covariates followed the definitions in Yonova-Doing *et al*. (2021)^10^ (described in supplementary table 6). In total, we assessed 826 phenotypes against 5,500 models and applied a multiple testing correction threshold of P < 1x10^-8^ to define significance (0.05/826*5500).

### Testing mechanisms of Breast Cancer and hypertension

To explore the mechanisms underlying associations between imputed mtDNA-encoded gene expression and binary disease traits, we conducted several downstream analyses. Specifically, for loci where a single nuclear variant appeared to drive the association, we performed both mediation and colocalization analyses. For mediation analysis, we assessed whether the effect of the nuclear genetic variant on the expression of the mtDNA-encoded gene was mediated through the expression of a nearby nuclear gene (i.e., in cis to the peak variant). We began by obtaining normalized expression data and covariates for individuals of European ancestry from the GTEx v8 portal, and matched these to the corresponding mtDNA-encoded gene expression data, generated as described above. For each nuclear gene within 1MB of the peak variant, we performed linear modeling to test for an association between variant genotype and nuclear gene expression, controlling for GTEx covariates. For nuclear genes showing a significant association (P<0.05), we then conducted mediation analysis using the *mediation* package in R, performing 1,000 simulations to estimate the indirect effect. Using the same GTEx data described above, we performed eQTL mapping for each nuclear gene within 1 Mb of the peak variant, using SNPs in the same 1 Mb window. We repeated this for the corresponding mtDNA-encoded gene. Separately, we performed logistic regression in the UK Biobank using PLINK to test the association between the same SNPs and the relevant binary disease trait, restricting to unrelated individuals of European ancestry and controlling for the same covariates as described above (age, sex, genotyping batch, and 10 genetic principal components). We then used the *coloc* package in R to perform colocalization analysis. For each gene in the 1 Mb window, we compared the eQTL summary statistics with GWAS summary statistics for the disease trait. This was done for both nuclear and mtDNA-encoded genes, allowing us to assess whether the same genetic signal drives both gene expression and disease risk.

### Comparison to age-related diseases

To evaluate the relationship between genetically predicted expression of mtDNA-encoded genes and common age-related diseases, we obtained GWAS summary statistics from five large-scale meta-analyses covering Parkinson’s disease^50^ (PD), Alzheimer’s disease^51^ (AD), coronary artery disease^52^ (CAD), atrial fibrillation^53^ (AF), and type 2 diabetes^54^ (T2D). Summary statistics for PD were sourced from the NDKP consortium (https://ndkp.hugeamp.org), T2D data from the DIAGRAM consortium (www.diagram-consortium.org) and the remaining traits from the GWAS Catalogue (www.ebi.ac.uk/gwas). For transcriptome-wide association testing, we used prediction models constructed with genetic variants filtered at a significance threshold of P < 1x10^-5^, in order to reduce the number of variants included and improve computational tractability. We applied all three modelling approaches (BLUP, Elastic Net, and LASSO) across all 49 GTEx tissue types where possible, resulting in a total of 2,121 distinct gene–tissue–model combinations. Summary statistic files were lifted over to the GRCh38 reference genome where required and converted to LD score format compatible with the FUSION pipeline. We then performed association testing using FUSION, comparing the weights of each mtDNA prediction model to the GWAS summary statistics using default parameters, except for adjusting the minimum required average imputation accuracy to 0.5 (--min_r2pred 0.5). To account for multiple testing across genes, tissues, models, and traits, we applied a Bonferroni correction. The significance threshold was set at P < 4.7 x 10^-6^, based on 0.05 divided by the total number of tests (2,121 models x 5 traits).

## Data availability

GTEx protected data are available through the database of Genotypes and Phenotypes (dbGaP) (accession no. phs000424.v8) and public-access data are available on the GTEx Portal (www.gtexportal.org). Genetic score files for mtDNA-encoded transcript abundance across 49 tissue types are available on Github (https://github.com/AJHodgkinson/mitoX).

## Code availability

All code used for analyses are available on Github (https://github.com/AJHodgkinson/mitoX).

## Acknowledgements

This work was supported by a Biotechnology and Biological Sciences Research Council (BBSRC) award to AH (BB/R006075/1). EH is supported by the BBSRC (BB/T008709/1). OP is supported by a Sir Henry Wellcome Postdoctoral Fellowship [222811/Z/21/Z]. The funders had no role in study design, data collection and analysis, decision to publish, or preparation of the manuscript. PFC is currently funded by a Wellcome Discovery Award (226653/Z/22/Z), a Wellcome Collaborative Award (224486/Z/21/Z), the Medical Research Council Mitochondrial Biology Unit (MC_UU_00028/7), and the Biological and Biotechnology Research Council (BB/Y003209/1), the Rosetrees Trust (PGL23/100048), and the LifeArc Centre to Treat Mitochondrial Diseases (LAC-TreatMito) under grant no. 10748. LifeArc is a charity registered in England and Wales under no. 1015243 and in Scotland under no. SC037861. His research is supported by the NIHR Cambridge Biomedical Research Centre (BRC-1215-20014). The views expressed are those of the author(s) and not necessarily those of the NIHR or the Department of Health and Social Care. The Genotype-Tissue Expression (GTEx) Project was supported by the Common Fund of the Office of the Director of the National Institutes of Health, and by NCI, NHGRI, NHLBI, NIDA, NIMH, and NINDS. The data used for the analyses described in this manuscript were obtained from dbGaP accession number phs000424.v8.p2. This work was facilitated by access to King’s Computational Research, Engineering and Technology Environment (CREATE) at King’s College London, UK. This research has been conducted using the UK Biobank Resource under application number 46360. We are grateful to the UK Biobank participants and team for their invaluable contributions. The project was approved by the UK Biobank and GTEx data access committees, and all analyses were conducted in accordance with the ethical approvals under which the data were originally collected. The Research Ethics Office of King’s College London gave ethical approval for this work (LRS-18/19-10868).

